# Social disparities in the first wave of COVID-19 infections in Germany: A county-scale explainable machine learning approach

**DOI:** 10.1101/2020.12.22.20248386

**Authors:** Gabriele Doblhammer, Constantin Reinke, Daniel Kreft

## Abstract

**Background:** Little is known about factors correlated with this geographic spread of the first wave of COVID-19 infections in Germany. Given the lack of individual-level socioeconomic information on COVID-19 cases, we resorted to an ecological study design, exploring regional correlates of COVID-19 diagnoses.

**Data and Method:** We used data from the Robert-Koch-Institute on COVID-19 diagnoses by sex, age (age groups: 0-4, 5-14, 15-34, 35-59, 60-79, 80+), county (NUTS3 region) differentiating five periods (initial phase: through 15 March; 1st lockdown period: 16 March to 31 March; 2nd lockdown period: from 1 April to 15 April; easing period: 16 April to 30 April; post-lockdown period: 1 May through 23 July). For each period we calculated age-standardized incidence of COVID-19 diagnoses on the county level, using the German age distribution from the year 2018. We characterized the regions by macro variables in nine domains: “Demography”, “Employment”, “Politics, religion, and education”, “Income”, “Settlement structure and environment”, “Health care”, “(structural) Poverty”, “Interrelationship with other regions”, and “Geography”. We trained gradient boosting models to predict the age-standardized incidence rates with the macro structures of the counties, and used SHAP values to characterize the 20 most prominent features in terms of negative/positive correlations with the outcome variable.

**Results:** The change in the age-standardized incidence rates over time is reflected in the changing importance of features as indicated by the mean SHAP values for the five periods. The first COVID-19 wave started as a disease in wealthy rural counties in southern Germany, and ventured into poorer urban and agricultural counties during the course of the first wave. The negative social gradient became more pronounced from the 2^nd^ lockdown period onwards, when wealthy counties appeared to be better protected. Population density per se does not appear to be a risk factor, and only in the post-lockdown period did connectedness become an important regional characteristic correlated with higher infections. Features related to economic and educational characteristics of the young population in a county played an important role at the beginning of the pandemic up to the 2^nd^ lockdown phase, as did features related to the population living in nursing homes; those related to international migration and a large proportion of foreigners living in a county became important in the post-lockdown period.

**Discussion:** In the absence of individual level data, explainable machine learning methods based on regional data may help to better understand the changing nature of the drivers of the pandemic. High mobility of high SES groups may drive the pandemic at the beginning of waves, while mitigation measures and beliefs about the seriousness of the pandemic as well as the compliance with mitigation measures put lower SES groups at higher risks later on.

## Introduction

Germany had comparatively few COVID-19 infections in the first wave (Vestergaard et al. 2020). There was a distinct south-north gradient with higher incidence in the south than the north (Scarpone et al. 2020), yet little is known about factors correlated with this geographic spread (Scarpone et al. 2020; Steiger et al. 2020). Cross-country studies showed that age structure (Esteve et al. 2020; Dowd et al. 2020; Dudel et al. 2020) had been shaping COVID-19 risk and in particular death from COVID-19 (Kulu und Dorey 2020), together with coresidence patterns (Esteve et al. 2020). Early studies from the start of the pandemic in China indicate occupational risk factors for health care workers, staff in tourism, retail and hospitality industry, transport and security workers, and construction workers, as well as staff attending international business meetings (Koh 2020). These professions do not follow an obvious social gradient. On the other hand, UK Biobank data showed that the risk of COVID-19 infection varied by ethnicity and socioeconomic status (Prats-Uribe et al. 2020), and regional studies revealed a higher risk for higher population density, higher proportion of Black and proportions of older adults (Liu et al. 2020). For New York City the heterogeneity in COVID-19 cases appeared to be largely driven by regional SES rather than mobility (Lamb et al. 2020).

One important factor that contributed to the spread of infection in Germany were hotspot events such as Ischgl in Austria, where Germans went for skiing holidays, and carnival festivities in South Germany (Felbermayr et al. 2020). A study using selected regional indicators related to economic, demographic, health and spatial characteristics of German regions did not find a relationship with income or unemployment rate, but did find a correlation with the number of employees in nursing professions (Ehlert 2020). Hospitalization data point towards a higher risk for the unemployed (Dragano et al. 2020).

No correlation was found with the density of built environment beyond the number of churches in a county (Scarpone et al. 2020). In a latter study labour market participation of the young appeared to be positively correlated with infections, i.e. low unemployment of persons aged 25 or younger, and high employment rates at ages 15 to 30 were correlated with higher infection rates (Scarpone et al. 2020).

None of this answers the question of whether a social gradient of COVID-19 infections existed for Germany. We would expect a gradient to be negative for the reasons pointed out for e.g. the UK (Patel et al. 2020):

First, lower SES groups live in more crowded environments, placing them at risk of lower respiratory tract infections, a major risk factor for severe COVID-19 infections. Poor housing conditions may also lower compliance with social distancing.

Second, many low SES groups do not have the opportunities to work from home, due to the nature of their occupations, thus they were less protected by lockdown measures.

Third, poverty and the related stress may not only increase the exposure to the virus, but also reduce the immune system’s ability to combat it.

Fourth, low SES groups may be less able to navigate the health care system, thus presenting at a later infection stage or being less able to communicate symptoms hindered by language barriers or attitudes towards ethnic minority groups.

Fifth, numerous risk factors are more common among low SES groups, such as hypertension, diabetes, lung and heart disease.

Finally, unequal access to information, differences in political preferences that may influence how information is processed, and attitudes toward risk may increase low SES infection risks (Weill et al. 2020).

In Germany, the expectation of a negative SES gradient is supported by after-lockdown hotspots in abattoirs and among fruit and vegetable short-term harvest workers (Neef 2020; Yapici 2020). This was attributed to the low temperatures and heavy physical work in abattoirs, combined with crowded and unhygienic living conditions. Being able to work from home office is socially stratified (Felbermayr et al. 2020), and risk factors of poor health were present in the majority of severe COVID-19 infections (Karagiannidis et al. 2020).

However, there are also reasons for a positive social gradient, at least at the beginning of the pandemic, as a study on social distancing responses in the US points out (Weill et al. 2020). Using mobility measures derived from mobile device location pings, they found a reversal of ordering of social distancing by income over the course of the pandemic: Wealthy areas went from most mobile before the pandemic to least mobile, while the poorest areas went from least mobile to most.

Given the lack of individual-level socioeconomic information on COVID-19 cases in Germany, we resorted to an ecological study design, exploring regional correlates of COVID-19 diagnoses. An ecological study design is hampered by the myriad of possible regional indicators, which often are highly correlated, and by the limited knowledge about the possible influence factors in relation to the time course of the pandemic. Using a data-driven approach, we applied methods of explainable machine learning to five distinct periods starting with the first COVID-19 case on 23 January 2020 in Bavaria through 23 July 2020. We used 166 different regional indicators on a county level and explored (1) to what extent the epidemiological information provided by the RKI, which is summarized below, is reflected in the regional indicators identified by the machine learning algorithms; and (2) whether there are indications of social gradients in the regional distribution of COVID-19 infections.

We hypothesized that the social gradient of infections changed over the course of the pandemic, which started with well-off (skiing) tourists returning from winter holidays in Austria and Italy, was further spread by carnival events in South Germany, but later affected workers in abattoirs and agriculture. However, it is unclear whether there was a general social gradient in terms of regions and, if so, if this positive or negative and when it occurred.

Given the reports about the large number of deaths in nursing homes in the most affected countries such as Spain, Italy, and the UK, we expect to find larger number of infections in regions with a large proportion of elderly who reside in nursing homes or who are dependent on care.

It is not clear if mobility between regions, in addition to the initial start of the disease, is of importance. The decline of mobility measured in terms of distance started on the weekend 14-15 March and by the end of March all federal states had agreed on common guidelines and regulations. The increase in mobility from mid-April onwards, however, did not result in increasing case numbers, but in further decrease (Bönisch et al. 2020). On the other hand, mobility had been the decisive factor at the start of the pandemic and it might still play an important role in spreading the disease out of hotspots. In New York City the subway system was critical for the spread of the disease from one district to another (Harris 2020), mobile phone geo-location was used to show how population outflows from Wuhan to other prefectures were related to the spread (Jia et al. 2020).

To explore these questions, we differentiated between five time periods. The first, the initial phase, covered the time span up through 15 March and was characterised by exponentially increasing infection diagnoses from the end of February onwards, with a reproduction value (R) well above 3. The second, covered the period from 16 March to 31 March and is referred to as the 1st lockdown period. First lockdown measures were introduced from 12th March onwards, with full lockdown starting 16th March. This lowered R to below 1.5. The third period, called the 2nd lockdown period, extended from 1 April to 15 April, during which R fell below 1 and reached a minimum of 0.5 around 15 April. Full lockdown was in place until around 19 April, when smaller shops (<800 m^2^), and zoos/parks started to reopen. The fourth period, referred to as the easing period, extends from 16 April to 30 April, with a gradual easing of lock-down measures in all counties. Finally, the fifth period covers 1 May through 23 July; a period in which R increased from roughly 0.3 up to levels fluctuating around 1, surging up in specific confined hotspots. Schools and shops started to re-open, masks became mandatory in public places such as shops, public transport, etc.. This is termed the post-lockdown period.

### Epidemiological Situation

The epidemiological situation in these five periods is displayed in terms of age-standardised infection rates in Figures 1a-1e and can be characterised as follows: In the **initial phase** through 15 March a total of 4,838 COVID-19 cases had been reported in all 16 federal states. Schools and day care centres were still open, as well as borders to neighbouring countries. The district of Heinsberg (North Rhine Westphalia) was a particularly affected area, and Tyrol in Austria had been determined to be a risk area as well (Robert Koch Institute 2020a).

**Figures 1a–1e;.**
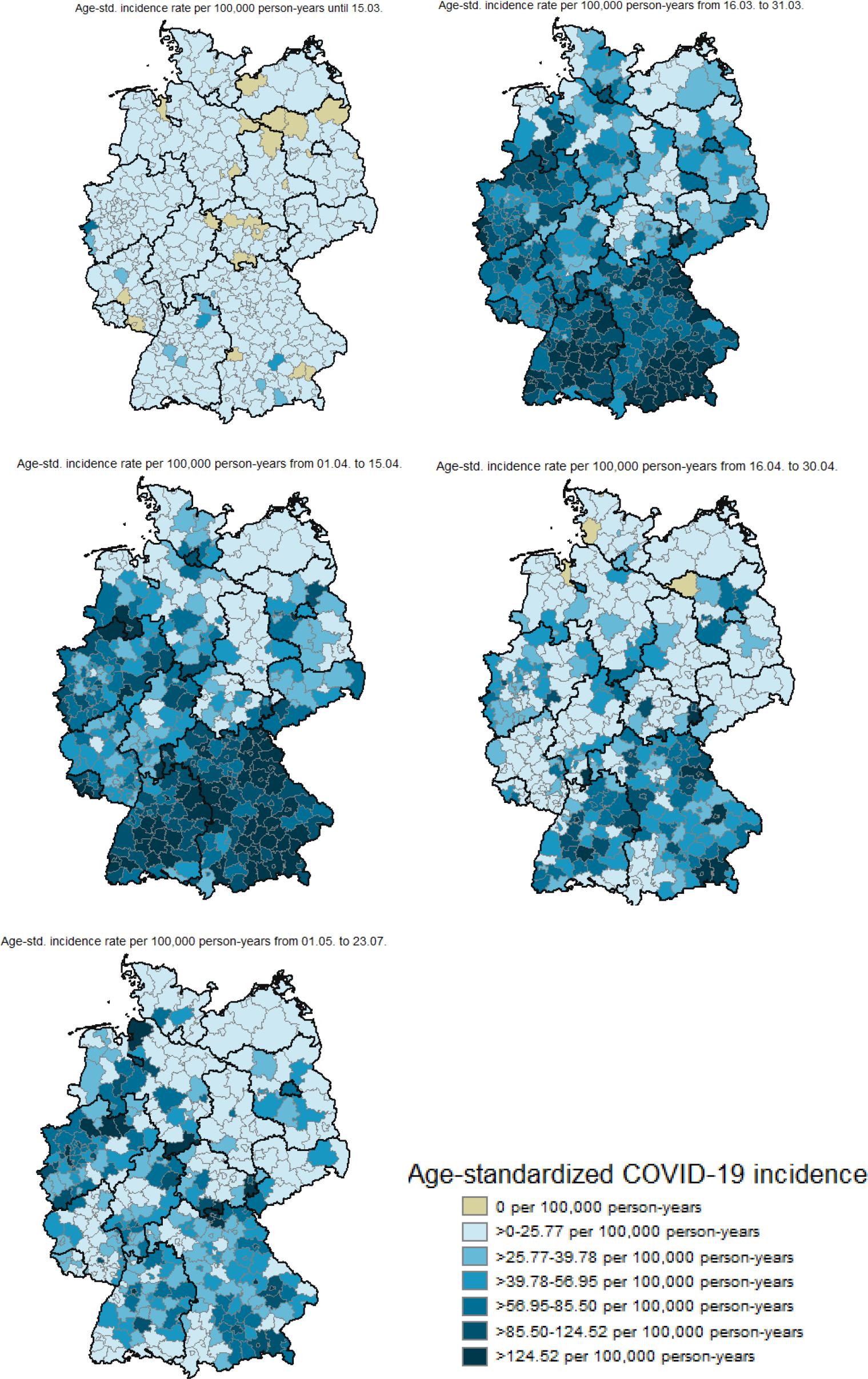
Age-standardized incidence of Covid-19 infections by period a) Initial Period: -15 March 2020; b) 1st Lockdown Period: 16 March - 31 March 2020; c) 2nd Lockdown Period: 1 April - 15 April; d) Easing Period: 16 April - 30 April 2020; e) Post-Lockdown Period: 1 May - 23 July 2020

By the end of the **1**^**st**^ **lockdown period** on 31 March 2020, 61,913 cases had been reported all over Germany. The incidence was highest in Hamburg (119 per 1000,000), Bavaria (113) and Baden-Württemberg (111). Most infected people were between 35 and 59 years old. As of 23 March gatherings of more than two persons (with the exception of families and household members) were prohibited, and restaurants and businesses with personal care were closed. A distance of 1.5 metres had to be maintained in public (Robert Koch Institute 2020b).

By the end of the **2**^nd^ **lockdown period** on 15^th^ April, 127,584 COVID-19 cases had been reported to the RKI. From 31 March onwards the RKI stopped identifying particular regions in Germany as heavily affected by COVID-19 because the virus had now spread throughout the country. The incidence continued to be highest in those regions that were already affected by the previous period, however Bavaria took the lead in infections with 262 cases. Nationwide, 68% cases were concentrated among the 15 and 59 year-olds, and only 18% of infections occurred in persons aged 70 years or older. An increasing number of outbreaks in nursing homes and hospitals were reported, with 6,058 cases among staff working in medical facilities. The number of tests had increased from 124,716 through week ten (2-8 March) to 360,139 in week 15 (6-13 April), with a positive rate increasing from 3.1% to 8.1% (Robert Koch Institute 2020c).

By the end of the **easing period** on 30 April, 159,119 cases had been reported to the RKI, with outbreaks in hospitals and nursing homes still ongoing. This was also true for the **post-lockdown period** with a total of 202,799 infections by 23 July. There were isolated clusters of COVID-19 infections in meat processing plants, as well as in facilities for asylum-seekers and refugees, nursing homes and hospitals, as well as in the context of family and religious events. In calendar week 29 (13-19 July) more than 530,000 tests were performed and only 0.6% of these were positive (Robert Koch Institute 2020d).

### Data

We used data from the Robert-Koch-Institute which provides information on COVID-19 diagnoses by sex, age (age groups: 0-4, 5-14, 15-34, 35-59, 60-79, 80+), and county (NUTS3 region). These were downloaded on 23 May 2020 through the publicly accessible NPGEO-DE platform (Robert Koch Institute und ESRI). Population size on county level was derived from the Regional database of the Statistical Offices of the Federation and the Länder at the end of the year 2018 (Statistische Ämter des Bundes und der Länder 2018). We calculated age-standardized incidence of COVID-19 diagnoses on the county level, using the German age distribution from the year 2018. We used age-standardized incidence rates because counties differ largely in their age distribution and age has been identified as one of the most important risk factors for severe COVID-19 infections.

Macro variables characterize counties in nine domains: “Demography”, “Employment”, “Politics, religion, and education”, “Income”, “Settlement structure and environment”, “Health care”, “(structural) Poverty”, “Interrelationship with other regions”, and “Geography”. The data stem from the INKAR (Indikatoren und Karten zur Raumund Stadtentwicklung) database (2020) of the Federal Institute for Research on Building, Urban Affairs and Spatial Development (BBSR), latitude and longitude were defined in terms of the centres of the county capitals. Air distance of the county centres to Ischgl was calculated by applying the equation: distance in km = sqrt(dx * dx + dy * dy) with dx = 111.3 * cos((lat1 + lat2) / 2 * 0.01745) * (lon1 - lon2) and dy = 111.3 * (lat1 - lat2), where lat1 and lon1 were the latitude and longitude of county 1 and lat2 and lon2 were the latitude and longitude of county 2. A dichotomous variable indicating more than 100 outbound commuters from the selected early hotspots Heinsberg, Tirschenreuth, Hohenlohekreit, Olpe, Aachen, Greiz, Saarbrücken, Potsdam, Coesfeld, Rosenheim, and Göttingen to the respective county stemmed from publicly available commuter flows from the Institute for Employment Research (IAB) for the year 2019, the proportion of Catholics in a county from the 2011 census. See Table 1 in Suppl. Material for the list of variables. All variables are numeric or dummies taking the values zero or one.

**Table 1:** Distribution of age-standardized COVID-19 incidence rates per 100,000 person-years by periods (n=401 counties, IQR interquartile range)

| Periods | Mean | SD | Min | 10% | 25% | 50% | 75% | 90% | Max | IQR |
| --- | --- | --- | --- | --- | --- | --- | --- | --- | --- | --- |
|  | per 100,000 person-years |  |  |  |  |  |  |  |  |  |
| Initial Period | 7.71 | 14.43 | 0.00 | 0.93 | 2.45 | 5.02 | 9.17 | 16.73 | 260.00 | 6.72 |
| 1st Lockdown Period | 79.46 | 64.12 | 2.71 | 23.95 | 40.31 | 63.81 | 99.63 | 146.69 | 671.13 | 59.33 |
| 2nd Lockdown Period | 79.01 | 72.68 | 3.61 | 17.81 | 34.00 | 56.94 | 107.51 | 158.03 | 721.38 | 73.50 |
| Easing Period | 35.32 | 34.21 | 0.00 | 6.50 | 11.89 | 25.52 | 47.31 | 76.35 | 223.53 | 35.42 |
| Post-Lockdown Period | 43.94 | 46.51 | 0.90 | 8.58 | 17.48 | 30.93 | 56.69 | 84.95 | 549.69 | 39.21 |

Initial Period: -15 March 2020; 1st Lockdown Period: 16 March -31 March 2020; 2nd Lockdown Period: 1 April - 15 April; Easing Period: 16 April - 30 April 2020; Post-Lockdown Period: 1 May - 23 July 2020

### Analysis strategy

As the exact nature of regional correlates on COVID19 infections is largely unknown, we opted for an exploratory data driven approach which does not impose our expectations on the model. We find this in machine learning approaches which have been successfully employed in predicting complex underlying relationships between input and response variables. For more complex methods based on decision trees, some studies have explored feature/variable importance where the predictors of a model are ranked by their importance in terms of the prediction performance of the model. However, ranking the variables does not capture the effects of the variables on individual predictions. (Lundberg und Lee 2017) proposed a unified framework for interpreting predictions named SHAP (SHapley Additive exPlanations) and we used the SHAP procedure to explore the importance of the 163 regional correlates with age-standardized COVID-19 incidence to gain insight into their association with individual projections, i.e. whether they are positively or negatively correlated with new COVID-19 cases.

First, we trained random forests and gradient boosting models to predict the age-standardized incidence rates with the macro structures of the counties, which are termed features (Figure 2). We did not split our data into a test and training sample because the data consist of the entire base population and we were interested in the best fitting model without having to consider overfitting to a specific sample. We used the random forest regressor from the Scikit-learn module in Python (Pedregosa et al. 2011) with 5000 trees. We kept all other hyperparameters at their default values. Gradient boosting models where trained using the CatBoostRegressor from the CatBoost algorithm (Prokhorenkova et al. 2018). This toolkit provides an efficient way to implement boosting models with high performance. We calculated the R^2^ and root mean squared (RMSE) errors to evaluate how well the models fit the data.

**Figure 2:**
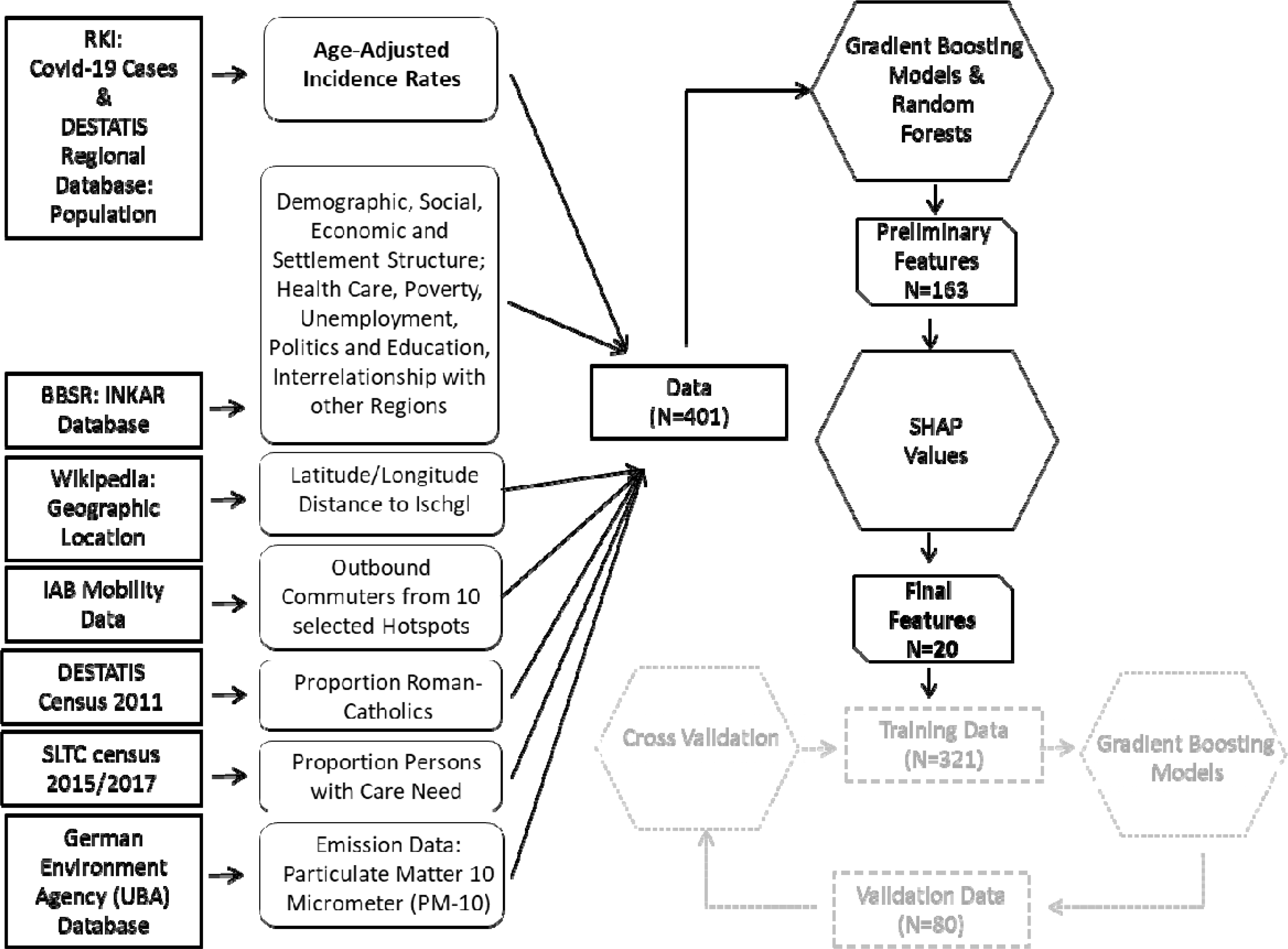
Analysis Flow

**Figures 3a – 3e:**
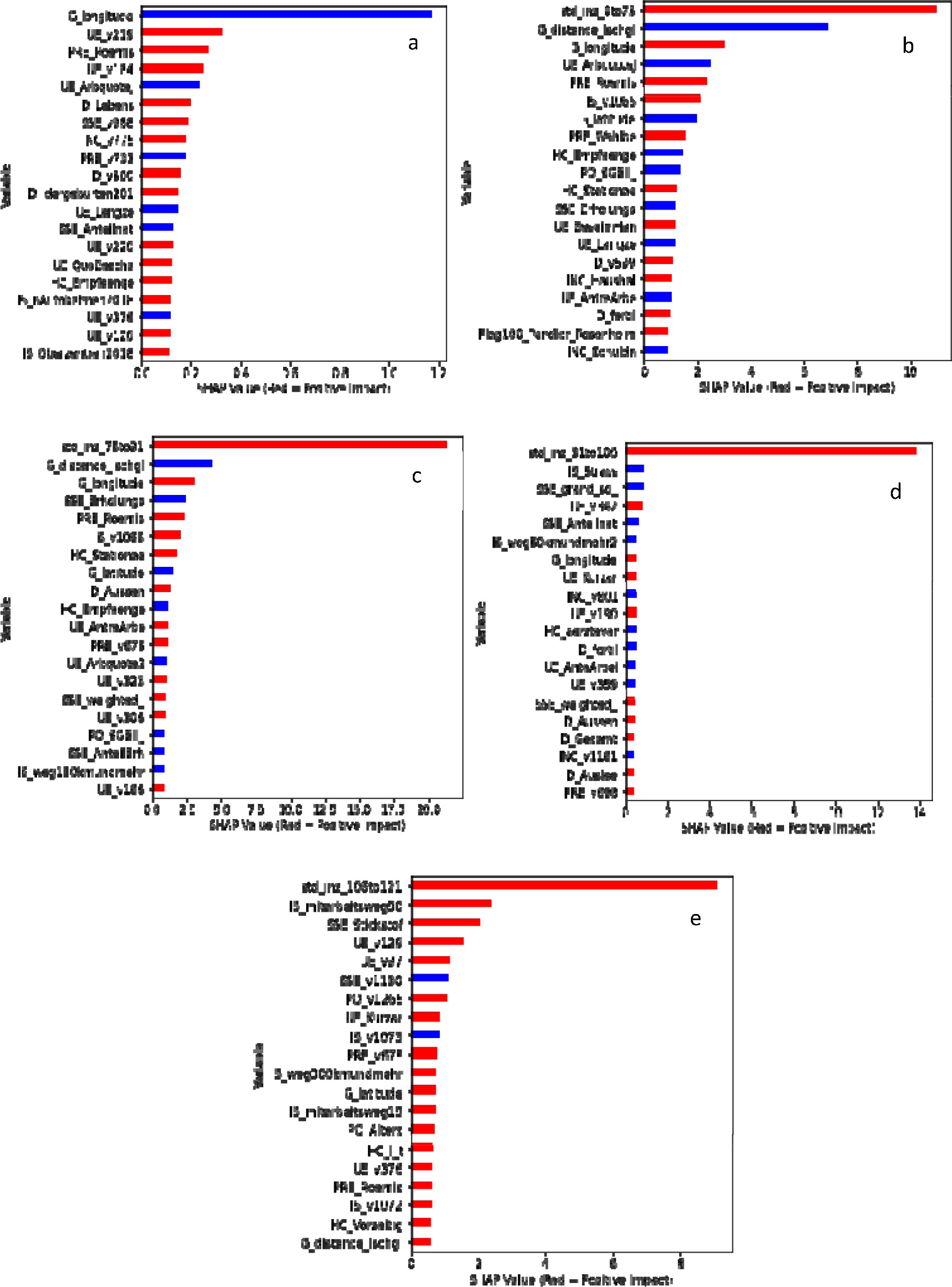
Mean SHAP values of the first 20 features identified by the Gradient Boosting Models by period Red: positive correlation with age-standardized incidence rates Blue: negative correlation with age-standardized incidence rates a) Initial Period: -15 March 2020; b) 1st Lockdown Period: 16 March -31 March 2020; c) 2nd Lockdown Period: 1 April - 15 April; d) Easing Period: 16 April - 30 April 2020; e) Post-Lockdown Period: 1 May - 23 July 2020 For legends see Suppl Table 1 and Suppl Tables 2a – 2e

Then we used SHAP values to characterize the 20 most prominent features in terms of negative/positive correlations with the outcome variable. We display the SHAP values in simplified SHAP summary plots, where the twenty features are ordered according to their importance. We display the mean over all regional SHAP values of the specific feature indicating whether a high/low value of the predicted outcome variable is correlated with a high/low value of the feature.

We categorized the associations into twelve categories depicting the correlation between the feature and the outcome: 1=positive SES gradient (SES high): higher incidence rates in high SES groups; 2=negative SES gradient (SES low): higher incidence in low SES groups; 3=urban/high density gradient (urban): higher incidence in urban/high density regions; 4=rural/low density gradient (rural): higher incidence in rural/low density regions; 5=poor health gradient (poor health): higher incidence associated with poor health; 6=good health gradient (good health): higher incidence associated with good health; 7= community’s connectedness low (connect low): higher incidence associated with low connectedness; 8=community’s connectedness high (connect high): higher incidence associated with high connectedness; 9=international migration high (migration high): higher incidence associated with high international migration; 10= Geography; 11= population characteristics; 12=other.

As a sensitivity analysis, we fit a second model for each period using only the 20 most prominent features from the first model. We calculated R^2^ and RMSE to evaluate how much variance in the age-standardized incidence rates is covered by using only the 20 most prominent features.

To evaluate the out-of-sample model performance, we followed the strategy of Scarpone et al. (2020) and applied k-fold random subsampling (Berrar 2019) using 20 folds. For each period we split the data at random to fit a model on a training set (80%) using the 20 most prominent features. This model was used to make predictions on a test set (20%) and to calculate the RMSE. Then a linear regression model was applied to explain the predictions by the actual response values from the test set. R^2^ from the linear regression model indicated how much variance from the actual response values could be explained by the predictions.

All analyses were performed using Python 3.8.3.

## Results

### 1. Age standardized COVID-19 incidence rates in the five periods

COVID-19 diagnosis rates revealed distinct geographic patterns that changed over time, as displayed in Table 1 and Figures 1a – 1e. In the initial period, only a few counties had high incidence rates, while 90 percent of all counties had rates lower than 16.73 cases per 100,000 person-years. The highest rates were registered in counties in South, Southwest and West Germany. The incidence steeply increased during the 1^st^ lockdown period, which was marked by profound clusters of high-incidence counties in South and North Bavaria, central Baden-Wurttemberg, and counties in North Rhine Westphalia. These clusters remained stable in the 2^nd^ lockdown period, but the maximum and the between-county range of the age-standardized rates increased further. The easing period showed the consequences of the lockdown period. In this period, the mean, median, and maximum rate and the between-county range declined. More than half of the counties had low and very low incidence rates (below 25.8 cases). Counties with the highest rates were still in Bavaria, Baden-Wurttemberg and North Rhine Westphalia. These patterns remained stable in the post-lockdown period with a slight increase in the cross-county mean, median, and the range of the incidence rates, but a steep increase in the maximum.

### 2. Model Fitting and Diagnostics

We decided to use the boosting models because in each period they outperformed the random forests in terms of accuracy (not shown). Using only the 20 most prominent features to fit boosting models resulted in nearly unchanged R^2^ scores but increased the RMSE scores (Table 2). This implied that the boosting algorithm produce well fitted models even when only a subset of the most prominent features is used. The out-of-sample performance varied over the periods. Especially the initial phase (period 1) as well as the post-lockdown period (period 5) showed a poor out-of-sample performance. For each period the descriptive statistics of outcome variable and the twenty most prominent features are presented in the in supplementary materials.

**Table 2:** R^2^ and RMSE scores of boosting models for all periods

|  | Initial<br>Period | 1st<br>Lockdown<br>Period | 2nd<br>Lockdown<br>Period | Easing<br>Period | Post-Lockdown<br>Period |
| --- | --- | --- | --- | --- | --- |
| $R^2$ first model (all features) | 0.9994 | 0.9994 | 0.9992 | 0.9990 | 0.9989 |
| RMSE first model (all features) | 0.3510 | 1.5513 | 1.9582 | 1.0441 | 1.5070 |
| $R^2$ second model (20 features) | 0.9967 | 0.9968 | 0.9969 | 0.9958 | 0.9914 |
| RMSE second model (20 features) | 0.8229 | 3.5810 | 4.0172 | 2.1960 | 4.2996 |
| $R^2$ second model (out-of-sample) | 0.1809 | 0.5687 | 0.6944 | 0.5739 | 0.2434 |
| RMSE second model (out-of-sample) | 12.7808 | 46.9289 | 40.4248 | 23.1244 | 41.9012 |

Initial Period: -15 March 2020; 1st Lockdown Period: 16 March -31 March 2020; 2nd Lockdown Period: 1 April - 15 April; Easing Period: 16 April - 30 April 2020; Post-Lockdown Period: 1 May - 23 July 2020

**Table 3:**
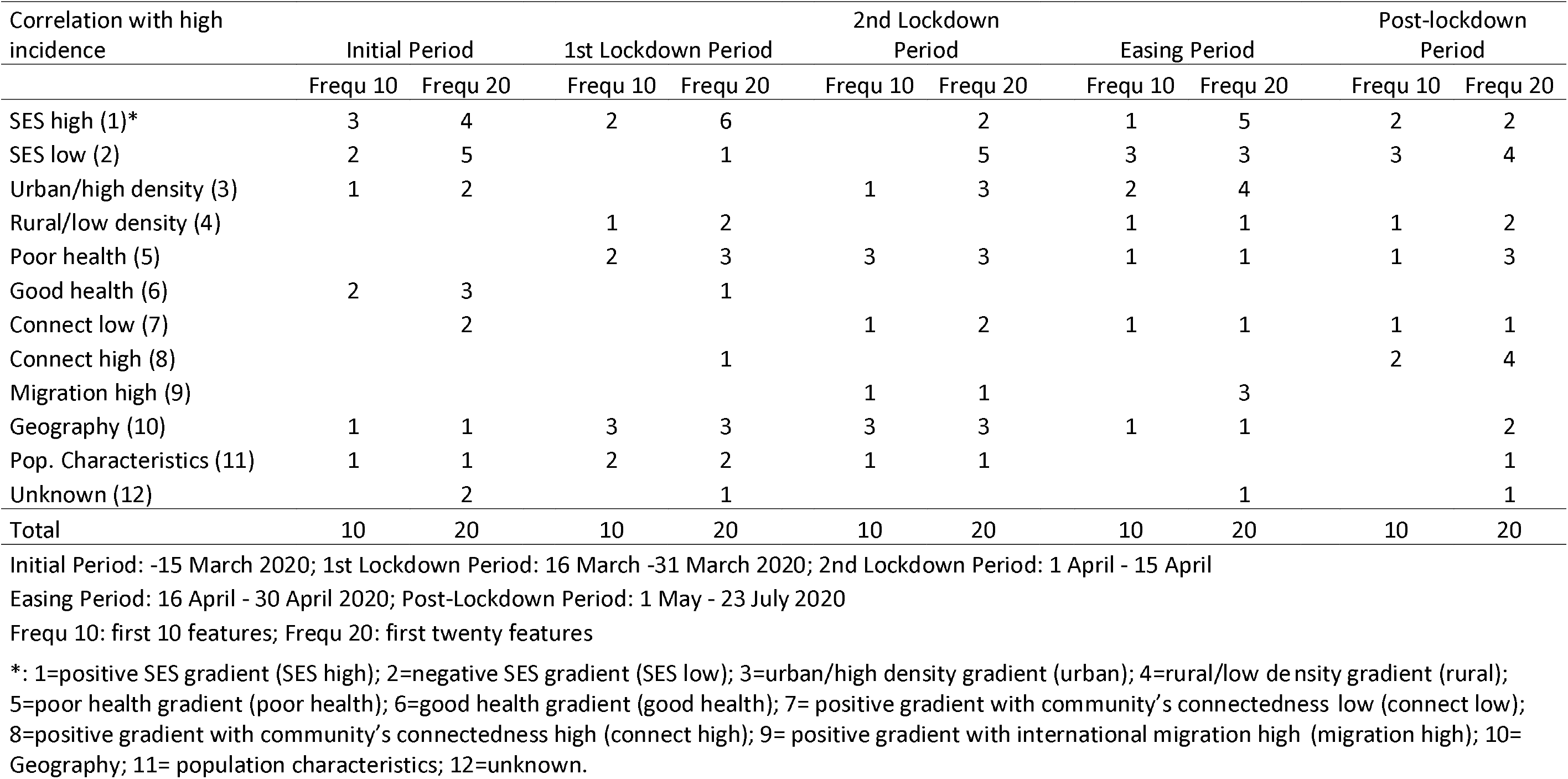
Number of features according to their type of correlation with COVID-19 incidence by period

| Correlation with high incidence | Initial Period |  | 1st Lockdown Period |  | 2nd Lockdown Period |  | Easing Period |  | Post-lockdown Period |  |
| --- | --- | --- | --- | --- | --- | --- | --- | --- | --- | --- |
|  | Frequ 10 | Frequ 20 | Frequ 10 | Frequ 20 | Frequ 10 | Frequ 20 | Frequ 10 | Frequ 20 | Frequ 10 | Frequ 20 |
| SES high (1)* | 3 | 4 | 2 | 6 |  | 2 | 1 | 5 | 2 | 2 |
| SES low (2) | 2 | 5 |  | 1 |  | 5 | 3 | 3 | 3 | 4 |
| Urban/high density (3) | 1 | 2 |  |  | 1 | 3 | 2 | 4 |  |  |
| Rural/low density (4) |  |  | 1 | 2 |  |  | 1 | 1 | 1 | 2 |
| Poor health (5) |  |  | 2 | 3 | 3 | 3 | 1 | 1 | 1 | 3 |
| Good health (6) | 2 | 3 |  | 1 |  |  |  |  |  |  |
| Connect low (7) |  | 2 |  |  | 1 | 2 | 1 | 1 | 1 | 1 |
| Connect high (8) |  |  |  | 1 |  |  |  |  | 2 | 4 |
| Migration high (9) |  |  |  |  | 1 | 1 |  | 3 |  |  |
| Geography (10) | 1 | 1 | 3 | 3 | 3 | 3 | 1 | 1 |  | 2 |
| Pop. Characteristics (11) | 1 | 1 | 2 | 2 | 1 | 1 |  |  |  | 1 |
| Unknown (12) |  | 2 |  | 1 |  |  |  | 1 |  | 1 |
| Total | 10 | 20 | 10 | 20 | 10 | 20 | 10 | 20 | 10 | 20 |

Initial Period: -15 March 2020; 1st Lockdown Period: 16 March -31 March 2020; 2nd Lockdown Period: 1 April - 15 April
Easing Period: 16 April - 30 April 2020; Post-Lockdown Period: 1 May - 23 July 2020
Frequ 10: first 10 features; Frequ 20: first twenty features
\*: 1=positive SES gradient (SES high); 2=negative SES gradient (SES low); 3=urban/high density gradient (urban); 4=rural/low density gradient (rural); 5=poor health gradient (poor health); 6=good health gradient (good health); 7= positive gradient with community's connectedness low (connect low); 8=positive gradient with community's connectedness high (connect high); 9= positive gradient with international migration high (migration high); 10= Geography; 11= population characteristics; 12=unknown.

### 3. Model results

The change in the age-standardized incidence rates over time is also reflected in the changing importance of features as indicated by the mean SHAP values for the five periods.

#### Period1: Initial phase

In the initial phase the most important feature was longitude (Figure 2a, Suppl. Table 2a), with incidence rates increasing the further west a county is situated. The second highest feature revealed a negative social gradient with higher incidence in counties with a higher “percentage of marginally employed persons in all persons 65+”; the third was related to regional population characteristics in terms of the “percentage of Roman-Catholics” with higher incidence rates. Among the first ten features there were three (3/10), which indicated a positive social gradient with higher incidence in wealthy counties (SES high), and two (2/10) with a negative SES gradient (SES low). Furthermore, there were two features with a positive gradient (2/10) with good health (good health) (Table 1). This pattern remained unchanged among the first twenty features (SES high: 4; SES low: 5; good health: 3). Urbanity/high population density was among the first ten features but was negatively correlated with incidence, which was reflected in the negative correlation for communities with a high connectivity among the first twenty.

In summary, in this period geographic location (west versus east) and a large population with Roman-Catholic denomination were the decisive factors. As expected, the latter was positively correlated with the outcome, displaying effects of the super-spreading events associated with carnival. We found higher incidence rates both in wealthy counties characterized by high SES and good health, as well as in poorer counties. Population density and high connectivity of regions were not driving the infections.

#### Period 2: 1^st^ Lockdown Period

Infections from the first period, and the distance to Ischgl became the two features with the highest importance, with declining incidence rates for increasing distance to Ischgl (Figure 2b, Suppl. Table 2b). Longitude and latitude now indicated higher incidence in the east and the south. The proportion of Roman-Catholics in a county still ranked fifth. Rurality and low population density still appeared to be associated with higher incidence rates. Wealthy counties were more affected with (6/20) features displaying a positive gradient with SES, and (2/20) with good health (Table 1). In summary, the geographical spread became more distinct with a focus in rural, lower populated areas. New infections were heavily influenced by the infections of the previous period in addition to the super-spreading events related to Carnival, as well as to Ischgl.

#### Period 3: 2^nd^ Lockdown Period

The most important features of the previous period are still present: previous incidence, distance to Ischgl, longitude, % Roman-Catholics. Low connectedness of a county was still correlated with high incidence, but in this period high population density and urbanity was positively associated with infections (Figure 2c, Suppl. Table 2c).With this shift to urban regions, poorer counties now had higher rates with (5/20) indicating a negative SES-gradient, and only (2/20) a positive. A total of (3/10) features pointed towards higher rates in counties with poor health, most notably towards counties with a large proportion of persons in inpatient long-term care among all persons in long-term care (Table 1). In summary, infections appeared to be concentrated more in urban and poorer counties with first indications of the vulnerability of counties with a large proportion of people living in nursing homes among those dependent on help.

#### Period 4: Easing Period

Infections were still spilling over from the previous period, while the next two highest ranking features indicated an urban/high density gradient (Figure 2d, Suppl. Table 2d). Still, poorer urban counties were affected with 3/10 features indicating a negative social, but 5/20 a positive urban gradient (Table 1). In summary, in the easing period the SES and urban/rural nature of infections continued to change with wealthy counties being obviously better protected than high SES counties and rural/low rural density counties better than urban/high density counties.

#### Period 5: Post-lockdown period

The trends of the easing period were re-enforced: poorer counties showed higher infection rates (4/20), which is also true for rural/less dense (2/20) and in particular agricultural areas as indicated by the positive correlation with the feature Nitrogen surplus (Figure 2e, Suppl. Table 2e). A county’s connectedness in terms of % outbound commuters/change in outbound commuters becomes an important feature ranking second and 4/20 features showing a positive correlation with infections (Table 1). In summary, while the negative SES gradient persisted, the infections moved back to rural/low density and agricultural areas, and transmission indicated by high connectedness between countries became an important pathway of spreading the disease.

## Discussion

Due to the lack of socio-economic information of COVID-19 infections in Germany, we resorted to an ecological study design with 166 regional variables on county-level possibly showing associations with infections. By using machine learning techniques we neither imposed our expectations on the analysis model, nor did we pre-select possible characteristics of the counties. We explored (1) whether the results reflected our knowledge about the epidemiological situation in the first wave of the pandemic as published in summary bulletins by the RKI; (2) whether indicators of SES can be identified, and (3) whether these changed over time.

Restricting our analysis to the first twenty risk factors identified by the variable importance we conclude that both social gradients, positive and negative, were present in COVID-19 infections right from the beginning, however, they changed over time. Distinguishing five time periods between February and mid-July 2020, we show that the first COVID-19 wave started as a disease in wealthy rural counties in southern Germany, and ventured into poorer urban and agricultural counties during the course of the first wave. The negative social gradient became more pronounced from the 2^nd^ lockdown period onwards, when wealthy counties appeared to be better protected. However, both negative and positive SES gradients were present over the full period. This course of the pandemic is consistent with findings from the US, where wealthier areas had higher mobility before the pandemic (Weill et al. 2020). In Germany this is reflected in the high feature importance of the distance to Ischgl, an international skiing-resort in the Alps, which was one of the hotspots of infections at the beginning of the pandemic. Return-mobility from the skiing resort may have contributed to thousands of Corona infections all over Europe (Felbermayr et al. 2020), with high SES groups being more likely having spent time there. The positive SES gradient remained strong until the 1^st^ lockdown period, while from the 2^nd^ lockdown period a strong negative gradient began to appear. Again, this is consistent with findings from US, where the wealthier areas decreased mobility significantly more than poorer areas (Weill et al. 2020). Finally, in the post-lockdown period features related to international migration and a large proportion of foreigners living in a county started to play an important role, again an indication of a negative social gradient with migrants being highly represented in occupations with system relevance and thus a higher potential exposition to the virus, such as cleaning workers, workers in food production, or nursing of elderly (DESTATIS 2020).

Superspreading events have been identified as an important driver of the pandemic, among them the carnival festivals in southern Germany (Streeck et al. 2020), which most probably are reflected in the feature “%Roman-Catholics” in a county and which is among the most important features until the 2^nd^ lockdown period. They contributed to the positive SES gradients because counties in southern Germany have higher SES and better health profiles. However, super-spreading events were also related to the emergence of the negative SES gradient, due to poor and little protected working and housing conditions in abattoirs and among agricultural workers, and to the spread of the disease in nursing homes even during the 1^st^ lockdown period (Althouse et al. 2020).

Population density per se does not appear to be a risk factor, as indicated by the lack of a clear urban-rural/density gradient, as well as by the negative gradient of a community’s connectedness measured in terms of time/distance to the next highway/train station. This lack of correlation with population density is supported by a regional analysis of COVID-19 prevalence in the US (Paul et al. 2020), as well as by Scarpone (2020) for Germany. It may be explained by the fact that cities have both the most healthiest population group, whose members benefit from better infrastructure and better access to health care, but also the least healthy groups, who have a higher burden of disease and lower life expectancy due to behavioural risk factors and exposure to environmental risk factors (Rydin et al. 2012). Only in the post-lockdown period did connectedness become an important regional characteristic correlated with higher infections, which may reflect the increase in mobility after the lockdown (Bönisch et al. 2020).

In our study, features related to economic and educational characteristics the young population in a county played an important role at the beginning of the pandemic up to the 2^nd^ lockdown phase. Infections among the young are less severe, often asymptotic, and frequently may not have been diagnosed in the first wave. Thus, our results suggest that as early as the first wave the young population may have considerably contributed to the spread of the virus. Again this is supported by (Paul et al. 2020), who concluded that the infections spread more easily among the elderly in regions where the population is younger. It is also supported by (Del Fava et al. 2020) who showed that social contacts decreased more rapidly among the older than the younger population.

We divided our periods into four two-week time-slots, which mainly reflect lockdown and easing measures, followed by a longer fifth period over more than 1.5 months, when infection rates were low. Our choice of period-duration is supported by (Dehning et al. 2020) in their change point analysis of the spread of COVID-19 in Germany, in which they found that change points in the spreading rate affected the confirmed case numbers with a delay of about two weeks. They observed three change points which are (1) the cancellation of large events with >1000 participants (around 9 March 2020), (2) the closing of schools, childcare centres, and most stores (in effect 16 March 2020), and (3) the contact ban and closing of all nonessential stores (in effect 23 March 2020). These three change points fall into the first two time periods of our study, where we observed a positive social gradient, and a positive gradient with good health. From our third period onwards, two weeks after the contact ban and the closing of nonessential stores, a strong negative social gradient emerged in our analysis, hence suggesting that these restrictions were more likely to protect high SES counties than low ones. This is consistent with a study of the work-from-home capacity in Germany before the pandemic (Alipour et al. 2020), which was lower among low-skilled and low-wage earners.

### Study Limitations

Our study is hampered by a series of limitations. Resorting to county level data does not only introduce the possibility of an ecological fallacy if results are interpreted on an individual rather than an aggregate level, but also the problem of the modifiable areal unit (Kirby et al. 2017). County level data might be too course but also too finely graded to detect important features driving the pandemic. Furthermore, they are limited to Germany and do not reflect if or how infections are acquired locally or internationally, with the exception of the variable “distance to Ischgl”.

True infection rates are unknown for COVID-19 because of asymptomatic individuals, regional eligibility criteria for testing leading to different testing rates, as well as differences in reporting of the local “Gesundheitsämter” to the RKI. To further complicate analyses, data from the RKI do not report the time of infection but rather of diagnosis, and by mid-April the date of the start of the illness was only known for 62% of the cases (an der Heiden und Hamouda 2020). Of these 50% were reported to the RKI within seven days, on 21 March it took 6.6 days, on 31 March it was 9.9, and in April it took 7.6 days. However, it has been shown that infected individuals are most contagious two to three days before symptoms start. In addition there was a strong weekday effect with lower numbers reported on weekends. Our 14-day time period averages over these various delays, yielding an average picture of infections in the time period. In addition we included information on infections in the previous period.

We did not include information on regional health profiles reflecting well known comorbidities of severe COVID-19 cases such as hypertension, diabetes, cardiac arrhythmia, renal failure, heart failure, and chronic pulmonary disease (Karagiannidis et al. 2020). These comorbidities are more common among persons with low SES and may be one pathway responsible for the negative social gradient observed in this study. However, we included general health measures such as (remaining) life expectancy and premature mortality, both of which are closely related to the chronic diseases mentioned above. Furthermore, we found positive gradients with both good and poor health measures as well as positive and negative SES gradients. This suggests that the relationship between chronic disease and (severe) COVID-19 infections is non-linear, and that mitigation measures play an important role.

Results from the use of machine learning algorithms to identify features and their importance depend on several factors, among them on the procedures implemented and this may produce spurious splits. We used both Random Forests (results available upon request) and Cat Boosting Algorithms, which led to similar conclusions. We relied on the latter because of better fit to the data in terms of R^2^ and RMSE. Nevertheless, one has to keep in mind that the SHAP values interpreted explain the model rather than the data. Our out-of-sample model fit was poor for both the initial and the post-lock down periods, which reflects the low number of incidence and the huge regional heterogeneity in infections at that time. It was high for periods with high incidence rates in a large number of counties, suggesting that our model prediction based on the first twenty features captures the dynamic of the pandemic well.

Our study shows that, in the absence of individual level data, explainable machine learning methods based on regional data may help to shed more light on the infection pathways of COVID-19 in Germany, and to better understand the changing nature of the drivers of the pandemic. Lessons for the second wave are that there appear to be no unique SES-drivers of the pandemic and dependent on the phase of the pandemic, different social groups are more or less affected. High mobility of high SES groups may drive the spread of the pandemic at the beginning of waves, while mitigation measures and beliefs about the seriousness of the pandemic as well as the compliance with mitigation measures (Galasso et al. 2020) may put lower SES groups at higher risks later on. To further substantiate this finding we urgently need individual level data on the socioeconomic background of COVID-19 patients (Khalatbari-Soltani et al. 2020) in Germany as well as internationally.

## Data Availability

All data referred to in the manuscript are publicly available.

https://npgeo-corona-npgeo-de.hub.arcgis.com/datasets/dd4580c810204019a7b8eb3e0b329dd6_0

https://www.regionalstatistik.de/genesis/online?operation=previous&levelindex=0&step=0&titel=Themen+%2F+Statistiken&levelid=1608606041212&acceptscookies=false#abreadcrumb

https://www.inkar.de/

https://www.umweltbundesamt.de/en/data

https://statistik.arbeitsagentur.de/DE/Navigation/Statistiken/Interaktive-Angebote/Pendleratlas/Pendleratlas-Nav.html

https://www.openstreetmap.de/karte.html

